# Bias in control selection associated with the use of rapid tests in influenza vaccine effectiveness studies

**DOI:** 10.1101/2024.11.16.24317422

**Authors:** Eero Poukka, Caitriona Murphy, Loretta Mak, Samuel M. S. Cheng, Malik Peiris, Tim K. Tsang, Sheena G. Sullivan, Benjamin J. Cowling

## Abstract

In test-negative design studies that use rapid tests to estimate influenza vaccine effectiveness (VE) a common concern is case/control misclassification due to imperfect test sensitivity and specificity. However, an imperfect test can also fail to exclude from the control group people that do not represent the source population, including people infected with other influenza types or other vaccine-preventable respiratory viruses for which vaccination status is correlated. We investigated these biases by comparing the effectiveness of seasonal 2023/24 influenza vaccination against influenza A and B based on PCR versus rapid test results, excluding controls who tested positive for SARS-CoV-2 or the other type of influenza. By PCR, VE against influenza A was 49% (95%CI 26–65%) after exclusion of PCR-confirmed influenza B and SARS-CoV-2 controls. Corresponding VE against influenza B was 65% (95%CI 35–81%). VE estimated by adjusting for COVID-19 vaccination status yielded similar estimates to the scenario that excluded SARS-CoV-2-positive controls. When case/control status and exclusions from test-negative controls were determined by rapid test, VE was reduced by 5–15 percentage points. Bias correction methods were able to reduce these discrepancies. When estimating VE from a test-negative study using rapid test results, methods to correct misclassification bias are recommended.

## Background

The test-negative study design (TND) is a commonly used approach for evaluating vaccine effectiveness (VE) and has long been used to monitor influenza VE in Hong Kong (1,2) and elsewhere (3–6). In a TND, participants are enrolled based on clinical criteria and tested for the pathogen of interest. The participants are classified as test-positive cases or test-negative controls according to the diagnostic test. The key premise in any case control study, including the TND, is that the enrollment of participants to the control group is independent of the exposure (vaccination), and controls represent the source population of the cases (7).

Imperfect diagnostic accuracy can lead to several potential issues. Consider the example of estimating influenza VE against influenza A(H1N1). A diagnostic test with imperfect sensitivity for A(H1N1) will give false negative results for some patients with A(H1N1), erroneously placing them in the test-negative control group. Under realistic assumptions of test sensitivity, this typically causes minimal bias in VE (8). Imperfect specificity may also be an issue, resulting in false positives being classified as test-positive cases, and potentially causing substantial bias (8). Some methods to correct this bias have been developed (9–11), one of them published by Endo et al (9). This method used multiple overimputation to correct VE estimates and provided unbiased estimates in simulation scenarios.

However, imperfect diagnostic accuracy causes other complications. In the analysis of VE against A(H1N1) discussed here, patients with influenza A(H3N2) or B should be excluded from the analysis because they do not have the same risk of infection by A(H1N1). Imperfect test sensitivity could result in failure to exclude some of these patients. In addition, as Doll et al. noted (12), health-seeking behavior that drives a person to be vaccinated against influenza may also drive them to be vaccinated against COVID-19, resulting in correlated vaccination status against the two diseases. In a TND, this can lead to increased enrollment of SARS-CoV-2 positive control participants who have not received either the influenza or COVID-19 vaccination and therefore do not represent the source population (Figure 1). Either adjusting for COVID-19 vaccination status or excluding SARS-CoV-2-positive controls have been proposed (12), and shown (13) to address this issue, but imperfect test accuracy for SARS-CoV-2 would affect such exclusions (Figure 1B).

**Figure 1.**
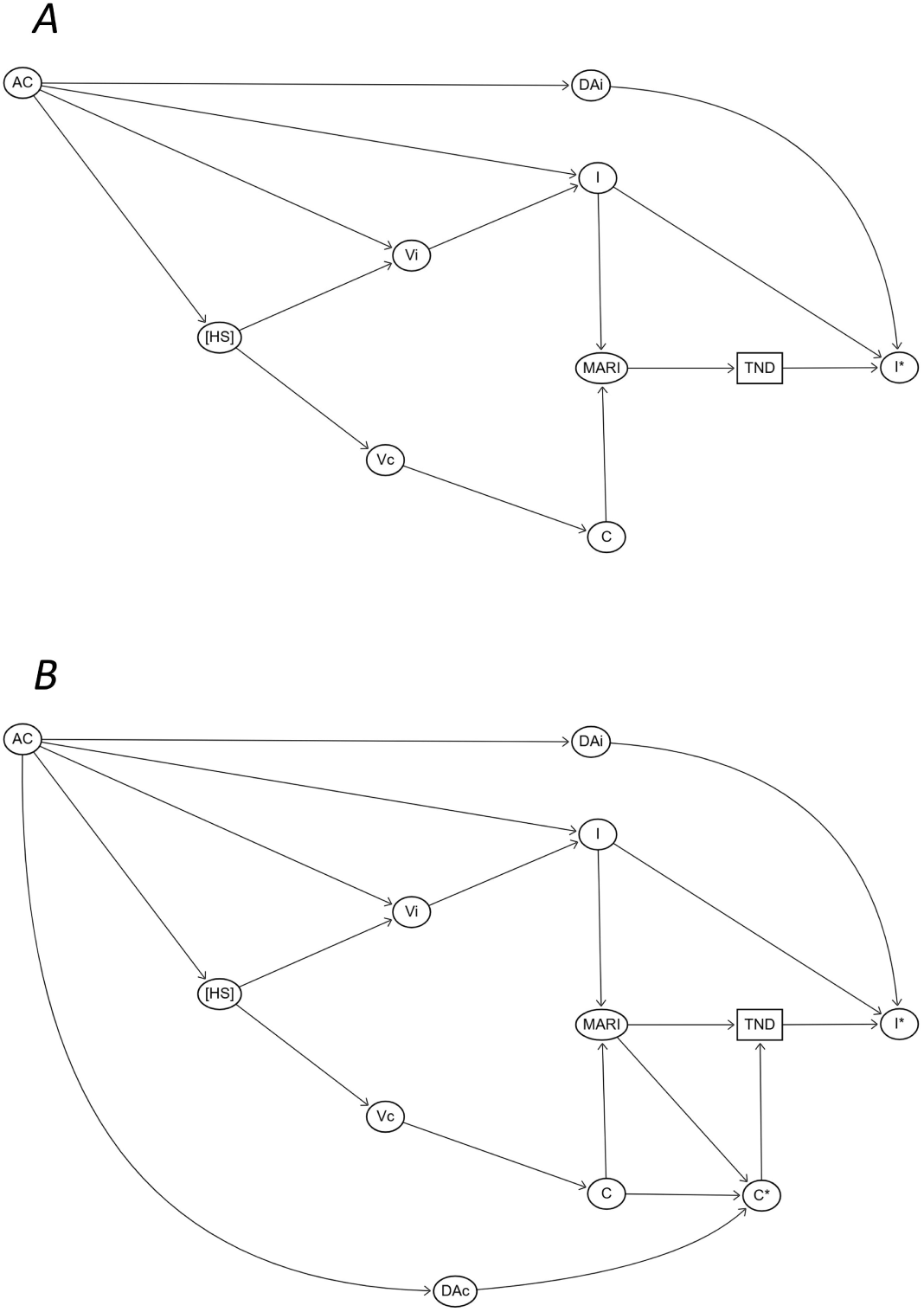
Directed acyclic graph showing the selection of participants in TND, including (A) or excluding (B) SARS-CoV-2 cases. HS = Health-seeking behaviour, Vi = Influenza vaccination, Vc = COVID-19 vaccination, AC = Age and chronic conditions, I = Infection due to influenza, C = Infection due to SARS-CoV-2, I* = Test detected influenza in a TND, C* = Test detected SARS-CoV-2, DAi = Diagnostic accuracy for influenza, DAc = Diagnostic accuracy for SARS-CoV-2, MARI = Medically attended acute respiratory infection, TND = Enrolment to a TND. In a TND evaluating VE against influenza, detection of influenza (I*) among the study participants influences case-control status in two ways: 1) other seasonal influenza types should be excluded from the controls due to cross protection of influenza vaccination, and 2) misclassification of true-positive cases as false-negative controls (6). The detection of influenza is dependent on diagnostic accuracy (DA_I_) and characteristics of the influenza infection (I). Another source of bias is correlation between the influenza (Vi) and COVID-19 vaccination (Vc) by health-seeking behaviour (HS) (12) which alters the selection of SARS-CoV-2 cases by influenza vaccination status (Vc ➔ C ➔ MARI ➔ TND). Notably, the correlation between V_I_ and V_C_ might be influenced by age and chronic conditions (AC) and can be expected to be more pronounced among priority groups, such as those with chronic illnesses and older adults. The correlation can be addressed by adjusting for COVID-19 vaccination (Vc in Figure 1A) or excluding positive SARS-CoV-2 cases (C* in Figure 1B) (12), with the latter depending on detection of the SARS-CoV-2 cases.

Our objective was to examine the potential bias introduced through use of rapid tests for estimation of influenza VE against influenza A and B in a TND among outpatients. We compared VE estimates obtained when PCR or rapid test results were used and evaluated the magnitude of bias resulting from the use of rapid tests to classify cases and controls and to make exclusions. We also applied the bias correction method of Endo et al (9) to assess whether valid VE estimates could be recovered after correcting for outcome misclassification. Finally, we compared the adjustment of COVID-19 vaccination and exclusions of SARS-CoV-2 from the controls to address the correlation between influenza and COVID-19 vaccination.

## Methods

### Study setting

In Hong Kong influenza vaccination is recommended for individuals aged ≥6 months, and almost all influenza vaccines administered each year are inactivated influenza vaccines (14). Hong Kong is located in a subtropical climate and influenza epidemics can occur at any time of the year, with winter peaks occurring most years and spring or summer peaks in some years (15). During the COVID-19 pandemic there was no influenza circulation in Hong Kong between March 2020 and February 2023 (16,17), but influenza activity resumed in March 2023 (18). Influenza circulated in Hong Kong in a series of epidemics from October 2023 through to August 2024, including spread of A(H3N2) from October to February, and A(H1N1) from March 2024 onwards (19).

### Study design and population

We conducted a TND study among outpatients aged at least 6 months of age and experiencing acute respiratory infection with at least two symptoms (fever ≥37.8°C, cough, sore throat, runny nose, headache, myalgia, and phlegm) starting within the preceding 72 hours. The study participants visited an outpatient clinic in Hong Kong between December 15, 2023, and August 13, 2024. Participants were interviewed by research staff using a standardized questionnaire and these responses were compared to medical records and vaccination certificates where possible. Swabs were collected for testing by PCR, and separate swabs were collected to conduct the rapid test “SARS-CoV-2 & Influenza A/B & RSV Antigen Kit” produced by *Goldsite Diagnostics Inc* (*Shenzhen, China*, Table S1) on-site. Since this rapid test was not widely available in the clinic before December 15, 2023, the study period began on this date. We excluded individuals that were not tested with this rapid test, had received seasonal influenza vaccination for 2023/24 within 13 days of the time of enrollment, or had incomplete background data (Figure S1).

### Specimens and outcomes

Pooled nasal and throat swabs were collected in a viral transport medium by trained research staff and tested by PCR for SARS-CoV-2, influenza A and B. In addition, separate nasal swabs were collected for rapid test to detect SARS-CoV-2, influenza A and B. If a rapid test gave an invalid result (control line did not appear), the test was repeated once. The outcomes of interest were influenza A and B confirmed by PCR or rapid test.

### Exposure

The exposure of interest was influenza vaccination for the 2023/24 season received at least 14 days prior to the time of enrollment. Those that had not received the seasonal 2023/24 influenza vaccination were classified as unexposed.

### Statistical analysis

We estimated VE against influenza A and B by PCR or rapid test using logistic regression. VE was calculated from the odds ratio (OR) comparing the odds of vaccination among influenza-positive cases versus influenza-negative controls, adjusted for confounders (VE = 1 – OR_adj_ x 100%). All models were adjusted for age, age-squared, sex, presence of diagnosed chronic medical conditions (cardiac, respiratory, hepatic, renal, hematological, immunological disease or diabetes ascertained by patient interview), receipt of seasonal influenza vaccination during the preceding season (2022/23) and calendar time (two-week bands).

To investigate the consequences of excluding influenza B and SARS-CoV-2 cases from the test-negative controls or alternatively addressing the correlation of influenza and COVID-19 vaccination by adjusting for COVID-19 vaccination status, we estimated VE against influenza A separately by PCR or rapid test using seven different analytic scenarios presented in Table 1.

**Table 1.** Seven scenarios for estimating VE against influenza A.

| Scenario | Influenza B |  | SARS-CoV-2 |  |  |
| --- | --- | --- | --- | --- | --- |
|  | PCR | Rapid test | PCR | Rapid test | Adjust for COVID-19 vaccination* |
| **1** | Exclude | NA | Exclude | NA | No |
| **2** | Exclude | NA | Include | NA | Yes |
| **3** | Exclude | NA | Include | NA | No |
| **4** | Include† | NA | Include | NA | No |
| **5** | NA | Exclude | NA | Exclude | No |
| **6** | NA | Exclude | NA | Include | Yes |
| **7** | NA | Exclude | NA | Include | No |

\*COVID-19 vaccine doses (0, 1–2 and 3 or more doses)
† No exclusions in the scenario.

When estimated against influenza A, scenarios 1 and 2 were considered the baseline analyses (the most valid estimates). For VE against influenza B the scenarios were similar, but the exclusions were instead based on influenza A positivity. As additional analyses we estimated the VE against influenza A separately among participants aged less than 18 years and participants aged 18 years or more. The age stratified analyses were not conducted against influenza B due to a small number of cases. We also estimated VE against influenza A during H3N2 (from December 15, 2023, until February 28, 2024) H1N1 (from March 1 until August 13, 2024) predominance periods to obtain estimates against specific subtype of influenza A.

To correct the VE estimates arising from imperfect diagnostic accuracy of rapid tests we used the multiple overimputation method introduced by Endo et al (9). The inputs for this method were the rapid test results, the predicted probability of a positive rapid test, the number of multiple overimputation iterations (chosen as 500) and estimates for sensitivity and specificity of the rapid test. The predicted probability of a positive rapid test for each study participant was estimated with the same logistic regression model used to estimate the OR between exposed and unexposed for different outcomes (i.e., the model used to estimate OR_adj_). For the sensitivity and specificity, we used estimates from the current study and in a separate analysis we used the values provided by the manufacturer (Table S1). The sensitivity and specificity of the rapid test were estimated from the study participants assuming PCR as gold standard (20). Separate sensitivity and specificity estimates were estimated by age groups (those aged less than 18 years and those aged 18 years or more) and these were similarly implemented to correct VE for the age group stratified analyses. The confidence intervals (CI) for sensitivity and specificity were estimated via the Clopper-Pearson intervals. The analyses were performed using R version 4.2.2 (R Foundation for Statistical Computing, Vienna, Austria).

### Ethical approval

The study received approval from the Institutional Review Board of the University of Hong Kong. Written informed consent was obtained for each participant and parental consent was obtained for participants below 18 years of age.

## Results

A total of 1,691 study participants were included in the study during the study period between December 2023 and August 2024 (Figure S1–S2). Among the participants 410 (24%) had PCR-confirmed influenza A and 178 (11%) had influenza B (Table 2). Among the influenza A cases, 184 (45%) were caused by influenza A(H1N1), 175 (43%) by A(H3N2), and 51 (12%) had influenza A which was not subtyped. Approximately half of the study participants were under 18 years of age (N=879, 52%) and 9% (N=154) had at least one chronic medical condition.

**Table 2.** Characteristics of cases and controls.

|  | Influenza A |  | Influenza B |  |
| --- | --- | --- | --- | --- |
|  | Cases | Controls | Cases | Controls |
| **N** | 410 | 1,281 | 178 | 1,513 |
| **Sex** |  |  |  |  |
| Male | 179 | 573 | 89 | 663 |
| Female | 231 | 708 | 89 | 850 |
| **Age groups (in years)** |  |  |  |  |
| 0–4 | 59 | 232 | 17 | 274 |
| 5–9 | 63 | 289 | 26 | 327 |
| 10–17 | 59 | 177 | 41 | 195 |
| 18–49 | 155 | 413 | 86 | 482 |
| 50+ | 74 | 170 | 9 | 235 |
| **Presence of chronic medical conditions\*** |  |  |  |  |
| Yes | 36 | 118 | 11 | 143 |
| No | 374 | 1,163 | 167 | 1,370 |
| **Influenza vaccination for season 2023/24** |  |  |  |  |
| Yes | 99 | 470 | 24 | 545 |
| No | 311 | 811 | 154 | 968 |
| **Previous season influenza vaccination (2022/23)** |  |  |  |  |
| Yes | 101 | 421 | 26 | 496 |
| No | 309 | 860 | 152 | 1,017 |
| **Number of COVID-19 vaccinations** |  |  |  |  |
| 0 | 80 | 253 | 20 | 313 |
| 1–2 | 98 | 354 | 53 | 399 |
| ≥3 | 232 | 674 | 105 | 801 |
| **Days since symptom onset** |  |  |  |  |
| One day | 248 | 684 | 80 | 852 |
| Two days | 66 | 201 | 32 | 235 |
| Three days | 96 | 396 | 66 | 426 |

\* Defined as cardiac, respiratory, hepatic, renal, hematological, immunological disease or diabetes ascertained by patient interview.

For influenza A, 97 study participants received false negative rapid test results and five received false positive rapid test results in comparison to PCR (Table S2). The estimated sensitivity of the rapid test for influenza A was therefore 76.3% (95% CI 71.9–80.4%; Table 3) while the specificity was 99.6% (95% CI 99.1–99.9%). For influenza B, there were 36 false negatives and one false positive (Table S3), corresponding to a sensitivity of 79.8% (95% CI 73.1–85.4%; Table 3) and specificity of 99.9% (95% CI 99.6–100%). For SARS-CoV-2, there were 24 false negative and no false positive rapid test results. The estimated sensitivity for SARS-CoV-2 was 88.2% (95% CI 82.9–92.3%) and specificity was 100% (95% CI 99.8–100%) compared to PCR gold standard. Among participants aged less than 18 years the sensitivity for influenza A was higher compared to those aged 18 years or more (85.1%; 95% CI 79.0–89.9% vs 69.4%; 95% CI 63.0–75.3%). Similarly, the sensitivity for influenza B seemed higher among participants aged less than 18 years (84.3%; 95% 74.7–91.4% vs 75.8%; 95% 65.9–84.0%). For SARS-CoV-2 the sensitivity was comparable across the age groups (Table 3).

**Table 3.** Sensitivity and specificity of Goldsite rapid test for influenza A, B and SARS-CoV-2 among study participants across age groups.

|  | Sensitivity (95% CI) | Specificity (95% CI) |
| --- | --- | --- |
| **Influenza A** |  |  |
| All participants (N=1,691) | 76.3% (71.9 – 80.4%) | 99.6% (99.1–99.9%) |
| 0–17-year-olds (N=879) | 85.1% (79.0–89.9%) | 99.7% (99.0–100%) |
| 18-year-olds or older (N=812) | 69.4% (63.0–75.3%) | 99.5% (98.5–99.9%) |
| **Influenza B** |  |  |
| All participants (N=1,691) | 79.8% (73.1–85.4%) | 99.9% (99.6–100%) |
| 0–17-year-olds (N=879) | 84.3% (74.7–91.4%) | 99.9% (99.3–100%) |
| 18-year-olds or older (N=812) | 75.8% (65.9–84.0%) | 100% (99.5–100%) |
| **SARS-CoV-2** |  |  |
| All participants (N=1,691) | 88.2% (82.9–92.3%) | 100% (99.8–100%) |
| 0–17-year-olds (N=879) | 88.5% (77.8–95.3%) | 100% (99.6–100%) |
| 18-year-olds or older (N=812) | 88.0% (81.5–92.3%) | 100% (99.5–100%) |
| *CI* | *=* | *Confidence interval* |

VE against influenza A was 49% (95% CI 26–65%, Table 4) when cases were confirmed by PCR and participants testing positive for influenza B or SARS-CoV-2 by PCR were excluded. VE was 47% (95% CI 24–63%) against influenza A by PCR when influenza B-positive controls were excluded by PCR and COVID-19 vaccination status was adjusted. With other scenarios the VE by PCR varied from 42% to 47% (see Table 4). When influenza A case status was confirmed by PCR and no control exclusions were made, VE was underestimated at 42% (95% CI 17–60%).

**Table 4.** Vaccine effectiveness (VE) against influenza A based on PCR or rapid test with 95% confidence interval.

| Scenario | VE by PCR<br>(95% CI) | VE by rapid test<br>(95% CI) | Bias-corrected VE by rapid test<br>(95% CI) * | Bias-corrected VE by rapid test<br>(95% CI) † |
| --- | --- | --- | --- | --- |
| 1. Exclusion of influenza B and SARS-CoV-2 by PCR (N=1,314) | 49% (26–65%) ‡ | 45% (19–64%) | 49% (30–63%) | 48% (29–62%) |
| 2. Exclusion of influenza B by PCR and adjusting for COVID-19 vaccination (N=1,513) | 47% (24–63%) ‡ | 44% (18–63%) | 48% (36–58%) | 47% (36–55%) |
| 3. Exclusion of influenza B by PCR (N=1,513) | 47% (25–64%) | 44% (18–63%) | 48% (22–65%) | 47% (28–60%) |
| 4. All study participants (N=1,691) | 42% (17–60%) | 39% (9–59%) | 42% (12–61%) | 41% (21–56%) |
| 5. Exclusion of influenza B and SARS-CoV-2 by rapid test (N=1,370) | 47% (23–63%) | 43% (16–62%) | 47% (27–61%) | 46% (26–60%) |
| 6. Exclusion of influenza B by rapid test and adjusting for COVID-19 vaccination (N=1,548) | 45% (21–62%) | 42% (15–61%) | 46% (34–55%) | 44% (34–53%) |
| 7. Exclusion of influenza B by rapid test (N=1,548) | 45% (21–62%) | 42% (1 –61%) | 45% (26–60%) | 44% (25–59%) |

\* Diagnostic accuracy estimates based on data from the study participants (sensitivity = 76.3% and specificity = 99.6%)
† Diagnostic accuracy estimates of manufacturer (sensitivity = 85.0% and specificity = 99.3%)
‡ These estimates should be considered the most valid in this table

VE against influenza A confirmed by rapid test was 43% (95% CI 16–62%, Table 4) when participants testing positive for influenza B or SARS-CoV-2 by rapid test were excluded. Corresponding VE was 42% (95% CI 15–61%) when influenza B was excluded by rapid test and COVID-19 vaccination were adjusted. VE against influenza A by rapid test was 39% (95% CI 9– 59%) when no control group exclusions were made. Using both PCR results for the exclusion of influenza B and SARS-CoV-2 and applying the bias correction method increased VE estimates (Table 4).

VE against influenza B confirmed by PCR was 65% (95% CI 35–81%) after exclusion of PCR-confirmed influenza A and SARS-CoV-2 patients from the control group (Table 5). Corresponding VE was 63% (95% CI 34–80%) when influenza A were excluded by PCR and COVID-19 vaccination was adjusted. If these controls were not excluded VE was 58% (95% CI 25–77%). VE against influenza B by the rapid test was 49% (95% CI 6–73%) when patients with influenza A and SARS-CoV-2 by rapid test were excluded from the control group. Excluding PCR-confirmed influenza A and SARS-CoV-2 and applying bias correction increased VE estimates. However, the best estimates by rapid test for influenza B were still approximately 10 percentage points lower compared to baseline analyses despite applying both exclusions by PCR and bias correction.

**Table 5.** Vaccine effectiveness (VE) against influenza B based on PCR or rapid test with 95% confidence interval.

|  | **VE based on PCR**<br>**(95% CI)** | **VE based on rapid**<br>**test (95% CI)** | **Bias-corrected VE based on**<br>**rapid test (95% CI) \*** | **Bias-corrected VE based on**<br>**rapid test (95% CI) †** |
| --- | --- | --- | --- | --- |
| 1. Exclusion of influenza A and SARS-CoV-2 by PCR (N=1,084) | 65% (35–81%)<sup>‡</sup> | 52% (10–75%) | 54% (13–75%) | 52% (12–74%) |
| 2. Exclusion of influenza A by PCR and adjusting for COVID-19 vaccination (N=1,281) | 63% (34–80%)<sup>‡</sup> | 51% (8–74%) | 53% (25–70%) | 51% (30–66%) |
| 3. Exclusion of influenza A by PCR (N=1,281) | 63% (33–80%) | 50% (7–74%) | 51% (10–74%) | 50% (9–73%) |
| 4. All study participants (N=1,691) | 58% (25–77%) | 44% (-3–70%) | 46% (0–70%) | 44% (-1–69%) |
| 5. Exclusion of influenza A and SARS-CoV-2 by rapid test (N=1,196) | 63% (32–80%) | 49% (6–73%) | 52% (10–74%) | 50% (8–72%) |
| 6. Exclusion of influenza A by rapid test and adjusting for COVID-19 vaccination (N=1,373) | 63% (32–80%) | 50% (8–74%) | 52% (25–69%) | 52% (26–69%) |
| 7. Exclusion of influenza A by rapid test (N=1,373) | 62% (31–79%) | 48% (5–73%) | 51% (9–73%) | 49% (8–72%) |

\* Diagnostic accuracy estimates based on data from the study participants (sensitivity = 79.8% and specificity = 99.9%)
† Diagnostic accuracy estimates of manufacturer (sensitivity = 95.0% and specificity = 100%)
‡ These estimates should be considered the most valid in this table

In the stratified analyses, VE against influenza A by PCR was among participants aged less than 18 years 55% (95% CI 30–72%, Table S4) when participants testing positive for influenza B or SARS-CoV-2 by PCR were excluded. Corresponding VE estimate among participants aged 18 years or more was 38% (95% CI -21–69%, Table S5). Among participants aged less than 18 years the VE against influenza A by rapid test was 50% (95% CI 21–69%) with exclusions of participants testing positive for influenza B or SARS-CoV-2 by rapid test. However, among participants aged 18 years or more the corresponding VE was 26% (95% CI -52–65%). Overall, VE against influenza A by PCR or rapid test appeared to be lower among participants aged 18 years or more and the difference between VE by PCR or rapid test appeared greater indicating more profound bias in this age group. However, statistical power was limited in the age group stratified analysis. During H3N2 and H1N1 periods, VE against influenza A by PCR were 59% (95% CI 31–76%) and 33% (-14–61%, Tables S6–7) when participants testing positive for influenza B or SARS-CoV-2 by PCR were excluded. Corresponding VE against influenza A by rapid test were 53% (95% CI 19–72%) and 26% (95% CI -32–59%) with exclusions of participants testing positive for influenza B or SARS-CoV-2 by rapid test.

Overall, adjusting for COVID-19 vaccination and exclusion of SARS-CoV-2 to address correlation between influenza and COVID-19 vaccination provided comparable VE estimates for influenza A and B with only a 2 percentage point difference between estimates and compatible precision (Tables 4–5). Similar findings were observed for influenza A in the age and influenza A subtype stratified analysis (Tables S4–7).

## Discussion

In this TND study among outpatients, the most valid VE estimates of seasonal 2023/24 vaccination for PCR-confirmed influenza A were approximately 50% while they were approximately 65% for PCR-confirmed influenza B from December 2023 to August 2024. When utilizing rapid test results, VE against influenza was approximately 5 and 15 percentage points lower for influenza A and B compared to the most valid VE estimates by PCR due to the bias caused by misclassification of cases and controls, and failure to exclude other types of influenza and SARS-CoV-2 cases from the controls. Both exclusion of influenza and SARS-CoV-2 by PCR (instead of rapid test) and the correction method by Endo et al (9) were able to mitigate the bias although estimates remained lower compared to VE against PCR-confirmed influenza. This bias was especially prominent for influenza B, with the best estimate being approximately 10 percentage points lower by rapid test compared to PCR, despite using the bias correction method and excluding PCR-confirmed influenza A and SARS-CoV-2.

Our VE estimates for PCR-confirmed influenza A were comparable to mid-season estimates among outpatient clinics in Canada during October 2023 – January 2024 (VE against PCR-confirmed influenza A 59%, 95% CI 48–68%) (21). Similarly, comparable VE estimates were reported in other studies conducted in 2023/24 among outpatients in the United Kingdom (22), the USA (3), China (23), and in a multinational European study (4). For influenza B, VE has ranged from 51% to 89% in different outpatient settings during 2023/24 seasons similar to our results (3,23). Gào et al. reported VE against laboratory-confirmed influenza virus infection by PCR or rapid test in a TND conducted in outpatient and emergency settings during the 2023/24 season and observed approximately a 15 to 20 percentage point drop in VE against influenza A or B when using rapid tests (24). Together with our results, these highlight the importance to address the bias related to use of rapid test results in a TND.

Notably, several recent TND studies evaluating VE against influenza or COVID-19 have used rapid tests results without bias correction (25–30). When estimating VE against respiratory viruses with rapid tests in a TND study two sources of bias should be considered: 1) misclassification of cases and controls, and 2) failure to exclude other types of influenza, SARS-CoV-2, and RSV cases (if RSV immunization has been introduced) from the test-negative control group. Currently no widely used method is available to correct these sources of bias and new correction methods would be useful in the future. The rapid test-based TND could have promising prospects and could be used to estimate VE against symptomatic disease in participatory cohorts limiting cost of a TND and complementing other studies estimating VE against medically-attended illness or hospitalization.

We also observed the diagnostic accuracy of the rapid test to be dependent on age group with higher sensitivity observed among those aged less than 18 years. Therefore, the magnitude of bias associated with use of rapid tests might be dependent on age and this was also indicated in the age group stratified analyses. Other factors that might influence diagnostic accuracy include viral load (31) and, for SARS-CoV-2, the patient’s COVID-19 vaccination status (32). There have also been some reports of reduced disease severity for influenza among vaccine recipients (33–37). Given that disease severity is correlated with viral load (38) and therefore the chance of a positive rapid test result, this could result in differential outcome misclassification by vaccination status, which in a case-control design can be difficult to correct (39). Future studies to confirm the importance of outcome misclassification should additionally consider these potential sources of bias.

We found that both methods to account for the correlation between influenza and COVID-19 vaccination – adjusting for COVID-19 vaccination or excluding SARS-CoV-2 positive controls – provided comparable VE estimates. Payne et al (13) similarly observed compatible estimates when estimating VE against COVID-19 with adjustment for influenza vaccination status or exclusion of influenza-positive controls, as did DeCuir et al. and Laniece et al (40,41). In our study, addressing the correlation increased VE estimates by around 5 percentage points, suggesting that the possible bias was relatively small. In Canada, for the same season the increase in VE observed for influenza A(H3N2) was just 1 percentage point (21). In the prior 2022/23 season, similar-magnitude increases of 1 to 6 percentage points were reported from Europe (42,43) and Canada (44). Slightly larger 5–10 percentage increase was observed in the 2021/22 season (45), and it is possible that the confounding caused by correlated vaccination status was stronger in the US setting where vaccination uptake for both COVID-19 and influenza vaccines was higher than in Hong Kong, Europe and Canada. This bias may diminish over time but could still be considerable in some study settings if the magnitude of the bias depends on factors such as seasonal activity of SARS-CoV-2 and influenza, vaccine protection, circulating variants and study population characteristics.

There are several potential limitations of our study. Residual confounding is possible, although we were able to control the most relevant confounders in our analysis (6,46). As another limitation, our sample was relatively small especially in the stratified analyses. Rapid tests were not able to distinguish influenza A(H3N2) and A(H1N1) and we therefore had to approximate VE by subtype using calendar time. Despite our attempts to verify the data collected, it is possible that the study might have included additional exposure and covariate misclassification, the influence of which were not explored. Moreover, we only had information of number of COVID-19 doses and the date of the last COVID-19 vaccination was missing for most of the participants and therefore we could not explore adjusting for time since the last COVID-19 vaccination as a confounder, which could have addressed the correlation between influenza and COVID-19 vaccination more adequately. Finally, we only estimated VE against medically-attended influenza among outpatients that sought care at a clinic, most of whom were young and did not have chronic conditions. The results might not be generalizable to other outcomes, such as influenza associated hospitalizations, or other populations, such as the elderly or individuals with conditions that increase the risk of severe influenza disease.

## Conclusions

In this outpatient TND we estimated VE was approximately 50% against PCR-confirmed influenza A and 65% against influenza B. We found that VE estimated using rapid test results was approximately 5 to 15 percentage points lower than VE by PCR. The reduced sensitivity of current rapid tests compared to PCR is not only an issue for correct classification of cases and controls, but also an issue for making appropriate exclusions from the control group. New methods for controlling misclassification bias could help adapt participatory cohorts for monitoring of VE against influenza, COVID-19, and RSV.

## Data Availability

All data produced in the present study are available upon reasonable request to the authors.

## Acknowledgments

This project was supported by the Health and Medical Research Fund, Health Bureau, The Government of the Hong Kong Special Administrative Region (grant no. INF-HKU-3), the Theme-based Research Scheme (Project No. T11-712/19-N) of the Research Grants Council of the Hong Kong SAR Government, and the National Institute of General Medical Sciences (grant no. R01 GM139926). EP is supported by Finnish Medical Foundation. BJC is supported by an RGC Senior Research Fellowship from the University Grants Committee of Hong Kong (grant number: HKU SRFS2021-7S03).

## Author contributions

All authors meet the ICMJE criteria for authorship. EP and BJC conceived the study. Data were collected by LM and SMSC. Data were analyzed by EP and CM. MP and BJC supervised the work. EP wrote the first draft of the manuscript. All authors provided critical review and revision of the text and approved the final version.

