## Supplementary material for "Bias in control selection associated with the use of rapid tests in influenza vaccine effectiveness studies"

**Table S1.** Detailed rapid test specification.

| | |
|---|---|
| Assay | SARS-CoV-2 & Influenza A/B & RSV Antigen Kit (Colloidal Gold) |
| Manufacturer | Goldsite Diagnostics Inc., Shenzhen, China |
| Country of Origin | P.R. China |
| Certification | CE-IVD |
| Swab Type | Nasal swab |
| Reported Sensitivity and specificity | SARS-CoV-2: 93.0%; 100%<br>Influenza A: 85.0%; 99.3%<br>Influenza B: 95.0%; 100%<br>RSV: 92.3%; 100% |
| Format | Cassette |
| Method | Colloidal Gold |
| Volume applied into card/cassette | 2 drops (~60µL) |
| Incubation | 15 minutes |
| Readout | Visual: colored band |
| Limit of detection | SARS-CoV-2: 49 TCID 50/mL Influenza A(H3N2): 4.0 x 104 TCID 50/mL<br>Influenza A(H1N1): 2.0 x 104 TCID 50/mL RSV: 1.6x10^4 TCID 50/mL |
| Cross reactivity against other human virus | The results showed no cross reactivity |

**Table S2.** Characteristics of participants according to rapid test of influenza A.

| | False negative | False positive | True negative | True positive |
|---|---|---|---|---|
| N | 97 | 5 | 1,276 | 313 |
| Sex | | | | |
| Male | 30 | 2 | 571 | 149 |
| Female | 67 | 3 | 705 | 164 |
| Age groups | | | | |
| 0–4 | 5 | 1 | 231 | 54 |
| 5–9 | 9 | 1 | 288 | 54 |
| 10–17 | 13 | 0 | 177 | 46 |
| 18–49 | 47 | 3 | 410 | 108 |
| 50+ | 23 | 0 | 170 | 51 |
| Comorbidities | | | | |
| Yes | 11 | 0 | 118 | 25 |
| No | 86 | 5 | 1,158 | 288 |
| Influenza vaccination for season 2023/24 | | | | |
| Yes | 22 | 1 | 496 | 77 |
| No | 75 | 4 | 807 | 236 |
| Previous season influenza vaccination (2022/23) | | | | |
| Yes | 24 | 0 | 421 | 77 |
| No | 73 | 5 | 855 | 236 |
| Number of COVID-19 vaccinations | | | | |
| 0 | 10 | 1 | 252 | 70 |
| 1–2 | 25 | 0 | 354 | 73 |
| ≥3 | 62 | 4 | 670 | 170 |
| Days since symptom onset | | | | |
| One days | 60 | 3 | 681 | 188 |
| Two days | 9 | 0 | 201 | 57 |
| Three days | 28 | 2 | 394 | 68 |

**Table S3.** Characteristics of participants according to rapid test of influenza B.

| | False negative | False positive | True negative | True positive |
|---|---|---|---|---|
| N | 36 | 1 | 1,512 | 142 |
| Sex | | | | |
| Male | 22 | 0 | 663 | 67 |
| Female | 14 | 1 | 849 | 75 |
| Age groups | | | | |
| 0–4 | 2 | 0 | 274 | 15 |
| 5–9 | 6 | 1 | 326 | 19 |
| 10–17 | 5 | 0 | 195 | 36 |
| 18–49 | 21 | 0 | 482 | 65 |
| 50+ | 2 | 0 | 235 | 7 |
| Presence of chronic medical conditions | | | | |
| Yes | 3 | 0 | 143 | 8 |
| No | 33 | 1 | 1,369 | 134 |
| Influenza vaccination for season 2023/24 | | | | |
| Yes | 1 | 1 | 544 | 23 |
| No | 35 | 0 | 968 | 119 |
| Previous season influenza vaccination (2022/23) | | | | |
| Yes | 3 | 1 | 495 | 23 |
| No | 33 | 0 | 1,017 | 119 |
| Number of COVID-19 vaccinations | | | | |
| 0 | 4 | 0 | 313 | 16 |
| 1–2 | 12 | 0 | 399 | 41 |
| ≥3 | 20 | 1 | 800 | 85 |
| Days since symptom onset | | | | |
| One day | 17 | 0 | 852 | 63 |
| Two days | 6 | 1 | 234 | 26 |
| Three days | 13 | 0 | 426 | 53 |

**Table S4.** Vaccine effectiveness (VE) against influenza A among participants aged less than 18 years by PCR or rapid test with 95% confidence interval.

| | **VE by PCR (95% CI)** | **VE by rapid test (95% CI)** | **Bias-corrected VE by rapid test (95% CI)*** | **Bias-corrected VE by rapid test (95% CI)†** |
|---|---|---|---|---|
| 1. Exclusion of influenza B and SARS-CoV-2 by PCR (N=738) ‡ | 55% (30–72%) | 53% (24–71%) | 55% (33–70%) | 55% (33–70%) |
| 2. Exclusion of influenza B by PCR and adjusting for COVID-19 vaccination (N=796) ‡ | 52% (25–70%) | 50% (20–69%) | 52% (41–62%) | 53% (42–62%) |
| 3. Exclusion of influenza B by PCR (N=796) | 52% (25–70%) | 51% (21–70%) | 53% (8–76%) | 54% (31–69%) |
| 4. All study participants (N=879) | 47% (17–67%) | 46% (13–67%) | 48% (14–69%) | 48% (22–65%) |
| 5. Exclusion of influenza B and SARS-CoV-2 by rapid test (N=755) | 53% (27–70%) | 50% (21–69%) | 53% (30–68%) | 53% (30–69%) |
| 6. Exclusion of influenza B by rapid test and adjusting for COVID-19 vaccination (N=808) | 50% (48–86%) | 48% (17–68%) | 50% (39–59%) | 51% (40–60%) |
| 7. Exclusion of influenza B by rapid test (N=808) | 50% (22–68%) | 49% (18–68%) | 51% (27–67%) | 51% (27–68%) |

VE = Vaccine effectiveness

* Diagnostic accuracy estimates based on data from the study participants (sensitivity = 85.1% and specificity = 99.7%)

† Diagnostic accuracy estimates of manufacturer (sensitivity = 85.0% and specificity = 99.3%)

‡ These estimates should be considered the most valid in this table.

Table S5. Vaccine effectiveness (VE) against influenza A among participants aged 18 years or more by PCR or rapid test with 95% confidence interval.

| | VE by PCR (95% CI) | VE by rapid test (95% CI) | Bias-corrected VE by rapid test (95% CI)* | Bias-corrected VE by rapid test (95% CI)† |
|---|---|---|---|---|
| 1. Exclusion of influenza B and SARS-CoV-2 by PCR (N=576) ‡ | 38% (-21–69%) | 26% (-53–65%) | 31% (-17–69%) | 28% (-16–55%) |
| 2. Exclusion of influenza B by PCR and adjusting for COVID-19 vaccination (N=717) ‡ | 41% (-10–69%) | 32% (-35–67%) | 37% (5–58%) | 35% (12–53%) |
| 3. Exclusion of influenza B by PCR (N=717) | 41% (-9–69%) | 31% (-38–66%) | 36% (-128–82%) | 34% (-4–58%) |
| 4. All study participants (N=812) | 37% (-19–67%) | 26% (-50–65%) | 29% (-51–66%) | 28% (-12–54%) |
| 5. Exclusion of influenza B and SARS-CoV-2 by rapid test (N=615) | 37% (-21–68%) | 26% (-52–65%) | 31% (-19–60%) | 27% (- 17–55%) |
| 6. Exclusion of influenza B by rapid test and adjusting for COVID-19 vaccination (N=740) | 39% (-14–68%) | 30% (-40–66%) | 34% (2–56%) | 33% (7–51%) |
| 7. Exclusion of influenza B by rapid test (N=740) | 39% (-14–68%) | 29% (-43–65%) | 34% (-11–60%) | 31% (-8–56%) |

VE = Vaccine effectiveness

* Diagnostic accuracy estimates based on data from the study participants (sensitivity = 69.4% and specificity = 99.5%)

† Diagnostic accuracy estimates of manufacturer (sensitivity = 85.0% and specificity = 99.3%)

‡ These estimates should be considered the most valid in this table.

**Table S6.** Vaccine effectiveness (VE) against influenza A by PCR or rapid test with 95% confidence interval during H3N2 predominance (December 2023 – February 2024).

| | **VE by PCR (95% CI)** | **VE by rapid test (95% CI)** | **Bias-corrected VE by rapid test (95% CI)*** | **Bias-corrected VE by rapid test (95% CI)†** |
|---|---|---|---|---|
| 1. Exclusion of influenza B and SARS-CoV-2 by PCR (N=751) ‡ | 59% (31–76%) | 54% (20–77%) | 58% (35–73%) | 57% (35–72%) |
| 2. Exclusion of influenza B by PCR and adjusting for COVID-19 vaccination (N=871) ‡ | 56% (27–74%) | 52% (18–73%) | 57% (43–67%) | 55% (42–66%) |
| 3. Exclusion of influenza B by PCR (N=871) | 58% (29–75%) | 53% (19–73%) | 57% (18–77%) | 56% (34–71%) |
| 4. All study participants (N=920) | 56% (27–74%) | 52% (17–73%) | 55% (19–75%) | 54% (31–70%) |
| 5. Exclusion of influenza B and SARS-CoV-2 by rapid test (N=778) | 58% (30–76%) | 53% (19–74%) | 57% (34–72%) | 57% (34–72%) |
| 6. Exclusion of influenza B by rapid test and adjusting for COVID-19 vaccination (N=882) | 55% (26–74%) | 51% (16–72%) | 55% (40–66%) | 54% (41–64%) |
| 7. Exclusion of influenza B by rapid test (N=882) | 57% (13–73%) | 52% (18–73%) | 56% (33–71%) | 55% (33–70%) |

VE = Vaccine effectiveness

* Diagnostic accuracy estimates based on data from the study participants (sensitivity = 76.3% and specificity = 99.6%)

† Diagnostic accuracy estimates of manufacturer (sensitivity = 85.0% and specificity = 99.3%)

‡ These estimates should be considered the most valid in this table.

Table S7. Vaccine effectiveness (VE) against influenza A by PCR or rapid test with 95% confidence interval during H1N1 predominance (March – August 2024).

| | **VE by PCR (95% CI)** | **VE by rapid test (95% CI)** | **Bias-corrected VE by rapid test (95% CI)*** | **Bias-corrected VE by rapid test (95% CI)†** |
|---|---|---|---|---|
| 1. Exclusion of influenza B and SARS-CoV-2 by PCR (N=563) ‡ | 33% (-14–61%) | 30% (-25–61%) | 33% (-8–59%) | 32% (-8–57%) |
| 2. Exclusion of influenza B by PCR and adjusting for COVID-19 vaccination (N=642) ‡ | 34% (-10–61%) | 32% (-21–62%) | 35% (11–52%) | 33% (12–48%) |
| 3. Exclusion of influenza B by PCR (N=642) | 33% (-12–61%) | 29% (-25–61%) | 33% (-24–64%) | 32% (-8–57%) |
| 4. All study participants (N=771) | 21% (-34–54%) | 17% (-48–54%) | 19% (-49–56%) | 18% (-26–47%) |
| 5. Exclusion of influenza B and SARS-CoV-2 by rapid test (N=) | 29% (-20–58%) | 26% (-32–59%) | 29% (-15–56%) | 27% (-16–54%) |
| 6. Exclusion of influenza B by rapid test and adjusting for COVID-19 vaccination (N=666) | 30% (-17–59%) | 27% (-29–60%) | 30% (5–48%) | 29% (9–45%) |
| 7. Exclusion of influenza B by rapid test (N=666) | 29% (-20–58%) | 25% (-32–58%) | 28% (-15–54%) | 27% (-13–53%) |

VE = Vaccine effectiveness

* Diagnostic accuracy estimates based on data from the study participants (sensitivity = 76.3% and specificity = 99.6%)

† Diagnostic accuracy estimates of manufacturer (sensitivity = 85.0% and specificity = 99.3%)

‡ These estimates should be considered the most valid in this table.

**Figure S1.** Study enrolment.

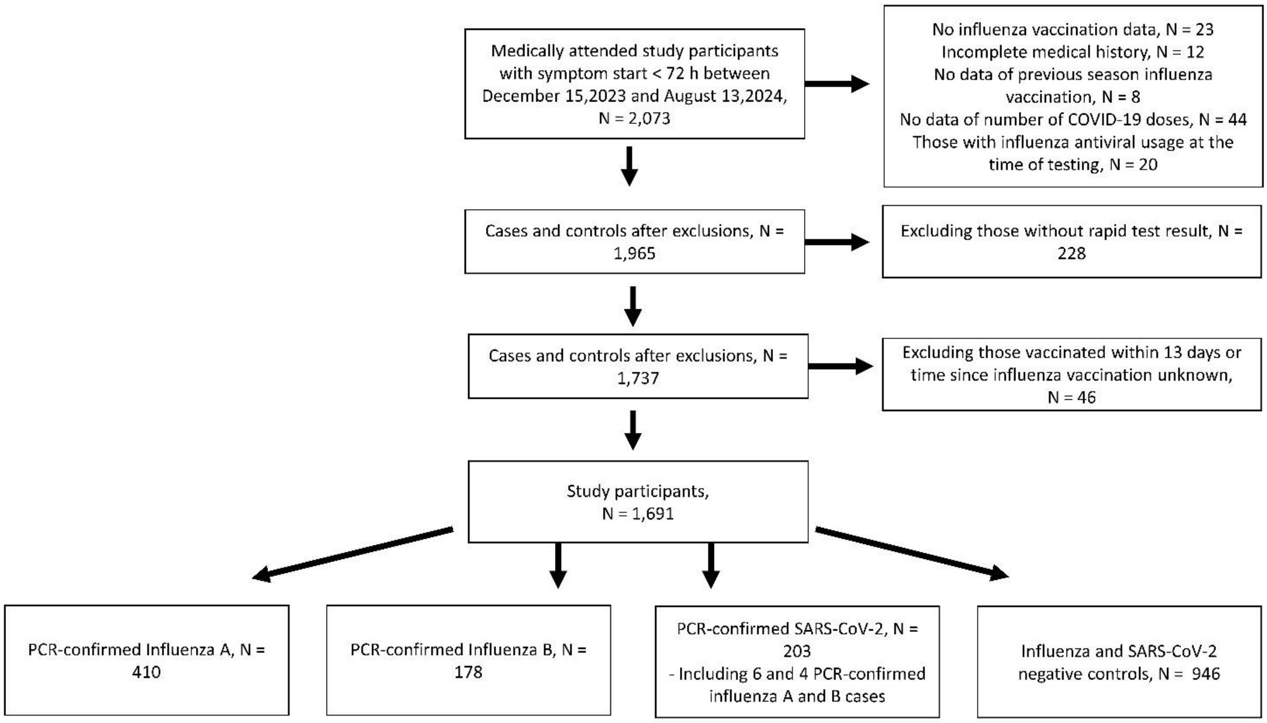

**Figure S2.** Influenza A and B cases over calendar time

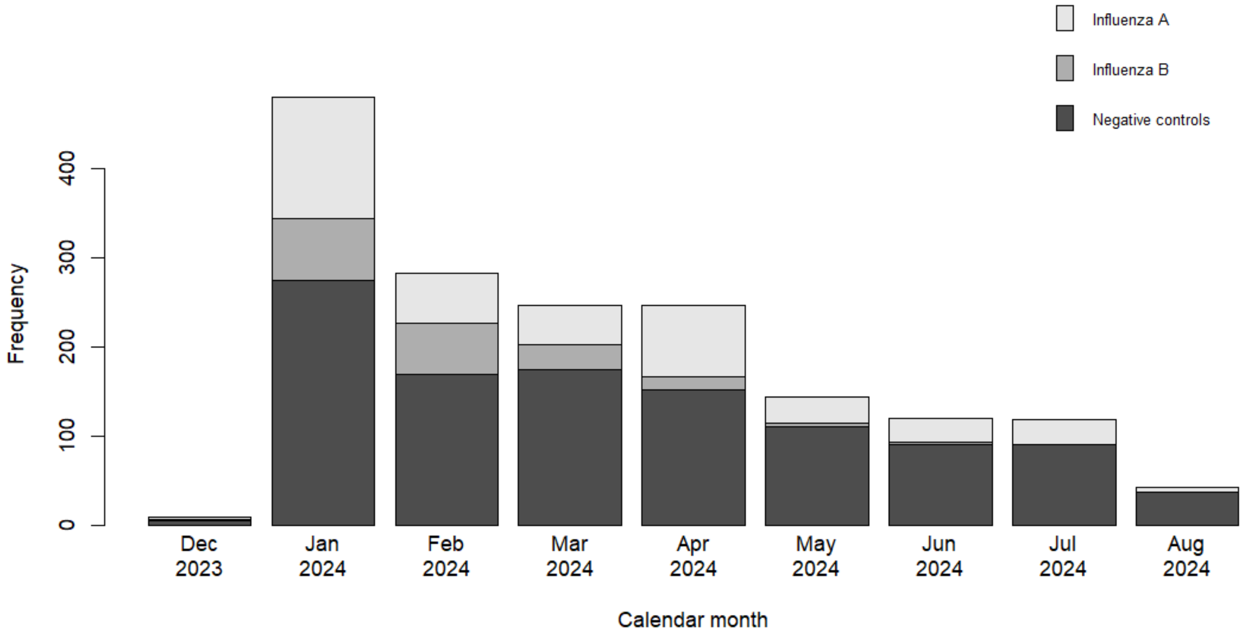
